# Duration of vaccine effectiveness against SARS-CoV2 infection, hospitalisation, and death in residents and staff of Long-Term Care Facilities (VIVALDI): a prospective cohort study, England, Dec 2020-Dec 2021

**DOI:** 10.1101/2022.03.09.22272098

**Authors:** Madhumita Shrotri, Maria Krutikov, Hadjer Nacer-Laidi, Borscha Azmi, Tom Palmer, Rebecca Giddings, Christopher Fuller, Aidan Irwin-Singer, Verity Baynton, Gokhan Tut, Paul Moss, Andrew Hayward, Andrew Copas, Laura Shallcross

**Affiliations:** UCL Institute of Health Informatics, London, UK; UCL Institute for Global Health, London, UK; Institute of Immunology and Immunotherapy, University of Birmingham, Birmingham, UK; Department of Health and Social Care, London, UK; UCL Institute of Epidemiology & Healthcare, London, UK; Health Data Research UK

## Abstract

**Background:** Long-term care facilities (LTCF) have been prioritised for vaccination, but data on potential waning of vaccine effectiveness (VE) and the impact of booster doses in this vulnerable population remains scarce.

**Methods:** We included residents and staff from 331 LTCFs enrolled in VIVALDI (ISRCTN 14447421), who underwent routine PCR testing between Dec 8, 2020 - Dec 11, 2021 in a Cox proportional hazards regression, estimating VE against SARS-CoV2 infection, COVID-19-related hospitalisation, and COVID-19-related death after 1-3 vaccine doses, stratifying by previous SARS-CoV2 exposure.

**Results:** For 15,518 older residents, VE declined from 50·7% (15·5, 71·3) to 17·2% (∼23·9, 44·6) against infection; from 85·4% (60·7, 94·.6) to 54·3% (26·2, 71·7) against hospitalisation; and from 94·4% (76·4, 98·7) to 62·8% (32·9, 79·4) against death, when comparing 2-12 weeks and ≥12 weeks after two doses. For 19,515 staff, VE against infection declined slightly from 50·3% (32·7, 63·3) to 42·1% 29·5, 52·4). High VE was restored following a third dose, with VE of 71·6% (53·5, 82·7) and 78·3% (70·1, 84·3) against infection and 89·9% (80·0, 94·6) and 95·8% (50·4, 99·6) against hospitalisation, for residents and staff respectively; and 97·5% (88·1, 99·5) against death for residents.

**Interpretation:** Substantial waning of VE is observed against all outcomes in residents from 12 weeks after a primary course of AstraZeneca or mRNA vaccines. Boosters restore protection, and maximise immunity across all outcomes. These findings demonstrate the importance of boosting and the need for ongoing surveillance of VE in this vulnerable cohort.

**Funding:** UK Government Department of Health and Social Care.

**Research in Context:** *Evidence before this study:* We searched MEDLINE and medRxiv for studies reporting vaccine effectiveness (VE) over time after two or three doses against SARS-CoV2 infection, COVID-19-related hospitalisation, or COVID-19-related death amongst staff or residents of long-term care facilities (LTCFs), that were published between Jan 1, 2020, and December 21, 2021. We used variations of the search terms “COVID-19” OR “SARS-CoV-2” AND “vaccine effectiveness” OR “vaccine efficacy” AND “care homes” OR “long term care facilities”. We identified 8 articles reporting two-dose data from LTCFs, including 1 peer-reviewed paper from Israel, 1 preprint from Denmark, 1 preprint from Norway, 1 peer-reviewed paper from France, two peer-reviewed papers from Spain, 1 peer-reviewed paper from the USA, and 1 preprint from England; however none of these studies examined waning of protection over time after two doses. Five studies (mRNA vaccines 3-4 weeks interval) reported short-term two-dose VE of 49-71% in residents, and 82-90% in staff. Two-dose VE was reported to be 75-88% against hospitalisation, 87-97% against death, and 86% against either outcome. An English study of residents (Pfizer or AstraZeneca, 8-12 week interval) reported 73% VE against infection and noted VE waning from 7 weeks after the first dose, but did not examine waning after the second dose. All of these studies were set prior to emergence of the Delta variant and did not examine waning of immunity due to short lengths of follow-up after Dose 2. Only one study (USA) compared Pfizer/Moderna two-dose VE against infection in LTCF residents before (67·5% [60·1-73·5%]) and during (53·1% [49·1-56·7%]) Delta variant predominance; however, authors could not access vaccination dates therefore did not account for any waning of immunity over time; they also did not examine any severe clinical outcomes. We identified only one correspondence piece from Israel (Pfizer 3-4 week interval) describing the benefit of a third ‘booster’ dose in LTCFs; it reported relative rate reductions of 71% for infection and 80%, for hospitalisation in the period after booster roll-out. However, individual-level VE estimates by time since vaccination were not reported, and adjustment for prior infection was not undertaken. Overall, there was a paucity of data on non-mRNA vaccines, waning of immunity over time after two doses, and VE following a third (booster) dose in LTCF populations, which we address in this study.

*Added value of this study:* We report findings from a prospective cohort study that includes 15,518 residents and 19,515 staff from 331 LTCFs across England, who underwent routine PCR testing 2-3 times per month, looking at SARS-CoV2 vaccine effectiveness over 12 months (Dec 8, 2020-Dec 11, 2021), which is the longest duration of follow-up of any study within this vulnerable cohort. We evaluated the effectiveness of first, second, and booster vaccine doses of AstraZeneca, Pfizer, and Moderna against infection, hospitalisation, and death over the 12 months when the Alpha and Delta variants were dominant. Our findings affirm that complete vaccination with two doses of AstraZeneca or mRNA vaccines offers moderate protection against infection, and high protection against severe clinical outcomes, however this protection declines over time, particularly for residents. A third booster dose of an mRNA vaccine restores, and indeed maximises, VE to 71·6% (53·5, 82·7) and 78·3% (70·1, 84·3) against infection, and 89·9% (80·0, 94·6) and 95·8% (50·4, 99·6) against hospitalisation, for residents and staff respectively, and to 97·5% (88·1, 99·5) against death for residents, with similar protection offered after the third dose irrespective of primary course type. This is the first study to examine and describe waning of immunity over a one-year period, as well as vaccine effectiveness of a booster dose, in a large cohort of LTCF staff and residents.

*Implications of all the available evidence:* Taken together, our findings indicate high short-term immunity against SARS-CoV2 infection and very high immunity against severe clinical outcomes of COVID-19 for LTCF residents and staff following vaccination. However substantial waning in vaccine-derived immunity is seen beyond 3 months, irrespective of vaccine type, suggesting the need for regular boosting to maintain protection in this vulnerable cohort. Although this analysis took place in the pre-Omicron period, these trends of waning immunity over time are likely to be generalisable across variants, carrying important implications for long-term vaccination policy in LTCFs. Ongoing surveillance in this vulnerable cohort remains crucial, in order to describe further changes in vaccine-induced immunity, particularly in the context of new variants.

## Introduction

Long-term care facility (LTCF) residents have been disproportionately affected by the COVID-19 pandemic, both in the UK and internationally,^1^ likely due to high levels of exposure to infection from staff or other residents within a closed setting, high levels of comorbidity and frailty, and age-related changes in immune function.^2^ Given these vulnerabilities, several policy measures were implemented in the UK including prioritisation of LTCF residents and staff for vaccination, routine testing of both residents and staff, and enhanced testing during outbreaks.^3,4^

In the UK, three vaccines have been deployed in LTCFs: Pfizer-BioNTech’s mRNA vaccine (tozinameran; Comirnaty), Oxford-AstraZeneca’s non-replicating viral-vectored vaccine (ChAdOx1-S; Vaxzevria) and Moderna’s mRNA vaccine (elasomeran; Spikevax), using an extended 8-12 week dosing interval. Clinical trials have demonstrated the efficacy of these vaccines against both infection and severe clinical outcomes in healthy adults, however older, more frail individuals are routinely excluded from trials^5^ therefore observational data has been crucial for understanding VE in this population.

Overall, vaccination has been highly effective in reducing infection and severe outcomes in LTCF residents and staff in the first 2-3 months.^6–12^ However, immunological data suggests that antibody levels start declining quickly following primary vaccination, with suggestions of more pronounced effects with older age.^13-15^ Recent VE estimates from the general population also suggest that protection against infection wanes over time.^16-18^ For these reasons, the extent and duration of protection afforded by vaccination in LTCF residents and staff is uncertain, particularly against newer variants such as Delta and Omicron which can partially evade the immune response.

Given emerging data on waning of vaccine-induced immunity, the JCVI recommended the use of a third ‘booster’ dose, to be given at least eight weeks after a primary course. LTCF residents and staff were prioritised for booster vaccination, which was rolled out from September 14, 2021.^19^ A key policy question is whether regular booster vaccines will continue to be required in this population, to mitigate waning of immunity and keep pace with the emergence of further new variants.

In this study we evaluate the effectiveness of first, second, and booster vaccine doses against infections, hospitalisations, and deaths amongst staff and residents of LTCFs in England, over the first 12 months following vaccination, covering the successive periods of Alpha and Delta variant predominance.

## Methods

### Study design and setting

The VIVALDI study is a prospective cohort study, investigating SARS-CoV-2 transmission, infection outcomes, and immunity in residents and staff of LTCFs from all regions in England.^20^ Since July 6, 2020, LTCFs in England have undertaken monthly routine asymptomatic SARS-CoV-2 testing using polymerase chain reaction (PCR)-based assays of nasopharyngeal swab specimens from residents. Staff undergo weekly testing using a combination of PCR and lateral flow devices (LFD). During the study period, all individuals with positive LFD results required confirmatory PCR testing with no routine retesting for 90 days. Vaccination of LTCF residents and staff in England commenced on Dec 8, 2020, initially with Pfizer, shortly followed by AstraZeneca (AZ). Primary courses consisted of homologous prime-boost regimens with an 8-12-week interval. Booster doses of Pfizer and Moderna were deployed in LTCFs from September 14 2021, during Delta variant predominance.

The analysis period started from Dec 8, 2020, the date of first vaccination in the VIVALDI cohort, and ended on Dec 11, 2021, the date from which Delta was superseded by Omicron as the predominant variant in England.^21^ As there was insufficient follow-up time to accurately account for the impact of the Omicron variant, which we expected to substantially alter VE estimates, we restricted this analysis to the pre-Omicron period. Residents and staff with at least two valid PCR test results, and at least one valid PCR result during the analysis period were eligible for inclusion in the analysis.

Ethical approval for the study was obtained from the South Central-Hampshire B Research Ethics Committee (20/SC/0238).

### Data extraction and linkage

We retrieved all PCR results and Cycle threshold values for PCR-positive samples from routine testing in LTCFs, and positive PCR results from clinical testing in hospitals through the UK’s COVID-19 Datastore.^22^ Void PCR tests were excluded from the analysis. Due to unreliability of symptoms data in the frail resident population and likelihood of pre-symptomatic positivity, symptoms data were not used. Ct values were only available for a subset of samples.

PCR results were linked to participants using pseudo-identifiers based on individuals’ unique UK National Health Service (NHS) numbers, which was also used to retrieve vaccination records (date, vaccine type, dose number) from the National Immunisation Management Service (NIMS); hospitalisation records (admission and discharge dates, ICD-10 diagnostic codes) from NHS England; and deaths data (date of death, ICD-10 codes for causes of death as per death certificate) from the Office for National Statistics through the COVID-19 Datastore. COVID-19-related hospitalisation was defined as an admission record associated with the ICD-10 code for COVID-19 (U071). COVID-19-related death was defined as inclusion of the ICD-10 code for COVID-19 on the death certificate.

A subset of participants also consented to serum sampling to detect IgG antibodies to the SARS-CoV2 Nucleocapsid protein using the Abbott ARCHITECT system (Abbott, Maidenhead, UK) SARS-CoV2 serological test results were also linked within the COVID-19 Datastore. A personal or nominated consultee was identified to act on behalf of residents who lacked the capacity to consent, and written, informed consent was given by those who were able. We combined positive PCR results, COVID-19-related hospitalisation records, and serological results from prior to the start of an individual’s risk period, into a binary variable indicating evidence of prior SARS-CoV2 exposure.

Participant PCR results could be linked to individual LTCFs using the unique provider identifier used by the national health and care regulator (Care Quality Commission), which is recorded at the point of testing. Data on bed capacity were retrieved from the NHS Capacity Tracker. Seven-day rolling rates of SARS-CoV2 incidence at the local authority level, produced by the UK Department of Health and Social Care,^22^ were used as a proxy for local infection pressure for each LTCF.

Palantir Technologies UK, under a general contract with the UK Government, provided the data platform for building the pseudonymised study dataset within the COVID-19 Datastore. The linked dataset was analysed in the University College London Data Safe Haven. A data privacy impact assessment was done for the VIVALDI study before the analysis.^23^ The legal basis for accessing data from staff and residents without informed consent was provided by the Control of Patient Information Regulations relating to COVID-19.

### Statistical analysis

We examined individual-level vaccine effectiveness against positive SARS-CoV2 PCRs, COVID-19-related hospitalisation, and COVID-19-related death, for residents aged ≥65 years, and for staff ≥18 years in VIVALDI LTCFs. The sample size for VIVALDI was based on the precision of estimates for antibody prevalence^20^ therefore, a-priori sample size calculations were not done for this analysis.

We used Cox proportional hazards models to derive adjusted hazard ratios (HRs) for the risk of each outcome of interest within the analysis periods. Vaccination status was included as a time-varying covariate, with the unvaccinated exposure group compared against exposure groups at 0-27 and ≥28 days following Dose 1, at 0-13, 14-83, and ≥84 days following Dose 2, and at any time from Dose 3. The same cohort contributed person-time at risk to unvaccinated and vaccinated exposure categories, with most individuals starting in the unvaccinated state and sequentially transitioning through vaccinated exposure states. Individuals entered the risk period on Dec 8, 2020 if they had at least one valid PCR result on or before that date; alternatively, they entered on the date of their first negative PCR test within the analysis period. Individuals with a positive PCR result within the 90 days prior to Dec 8, 2020, entered the risk period from the 91st day following their positive test. Individuals exited the risk period at the earliest of: the outcome of interest, their last valid PCR test (proxy for leaving the LTCF), or the end of analysis period on Dec 11, 2021. The baseline hazard was defined over calendar time. 95% CIs were calculated using robust SEs to account for dependence of infection events within LTCFs.

The primary analysis examined overall VE irrespective of vaccine type, stratified by evidence of SARS-CoV2 exposure prior to entering the risk period, using the unvaccinated unexposed group as the comparator. We adjusted for male sex (as a binary variable), age (as a cubic spline term), previous SARS-CoV2 exposure (as a binary variable), LTCF size expressed as total number of beds (as a linear term), local SARS-CoV2 incidence expressed as 7-day rolling rate per 100 population (as a linear term).

Secondary analyses stratified VE by first-dose vaccine type (Pfizer or Moderna [mRNA] vs AstraZeneca [AZ]). A sensitivity analysis of VE against infection censored from the point of Delta predominance (24th May 2021 onwards)^30^ to look at Dose 1 and early Dose 2 VE prior to Delta.

We calculated vaccine effectiveness estimates as 100□×□(1–adjusted HR) and 95% CIs for vaccine effectiveness estimates as 100□×□(1–upper and lower bounds of 95% CI for adjusted HR). We used two-tailed *t* tests to estimate the difference in mean Ct values of PCR-positive infections between groups, combining residents and staff.

All statistical analyses were conducted using STATA 16.0.

## Results

A total of 15,518 residents with a median age of 87 years (IQR 80, 92) of whom 68·3% (n=10,579) were female, and 19,515 staff with a median age of 45 years (32, 56) and of whom 84·5% (n=16,498) were female, were included in the analysis (Fig 1, Table 1). 11·5% (n=1,780) residents and 6·6% (n=1,281) staff had evidence of prior infection. The analysis period started from Dec 8, 2020, and ended on Dec 11, 2021, covering the emergence of Alpha, Delta, and Omicron variants and the periods of Alpha and Delta predominance (Fig 2a). During the analysis period, there were on average 2 (SE 0·024) and 3·11 (SE 0·017) valid PCR results per month-at-risk and a total of 129,965 and 373,666 results, for residents and staff respectively. The median follow-up time was 224 days (IQR 59,349) and 239 days (79, 357) in total and 221 days (117, 247) and 173 days (35, 245) after Dose 2, for residents and staff respectively.

**Table 1.**
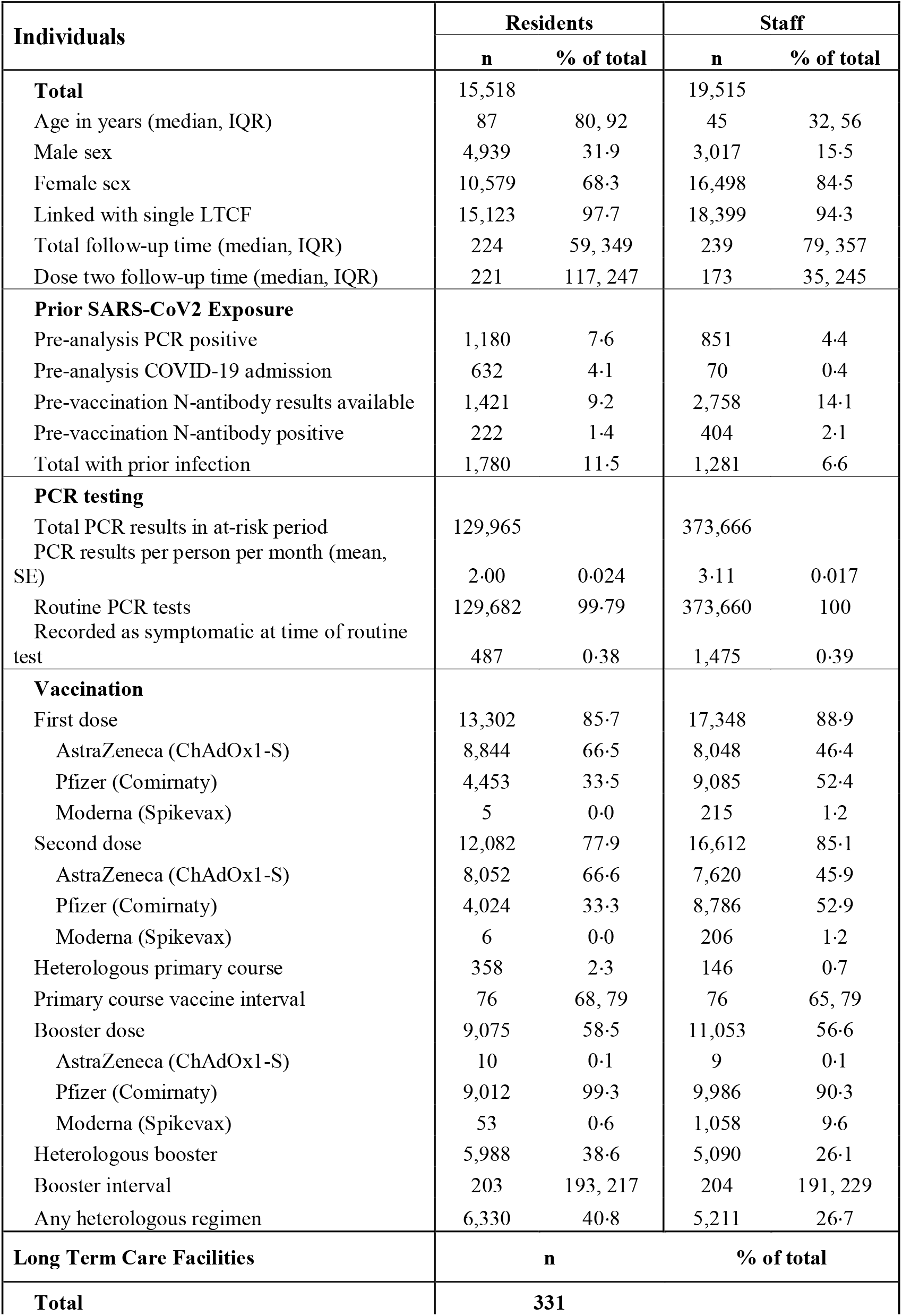
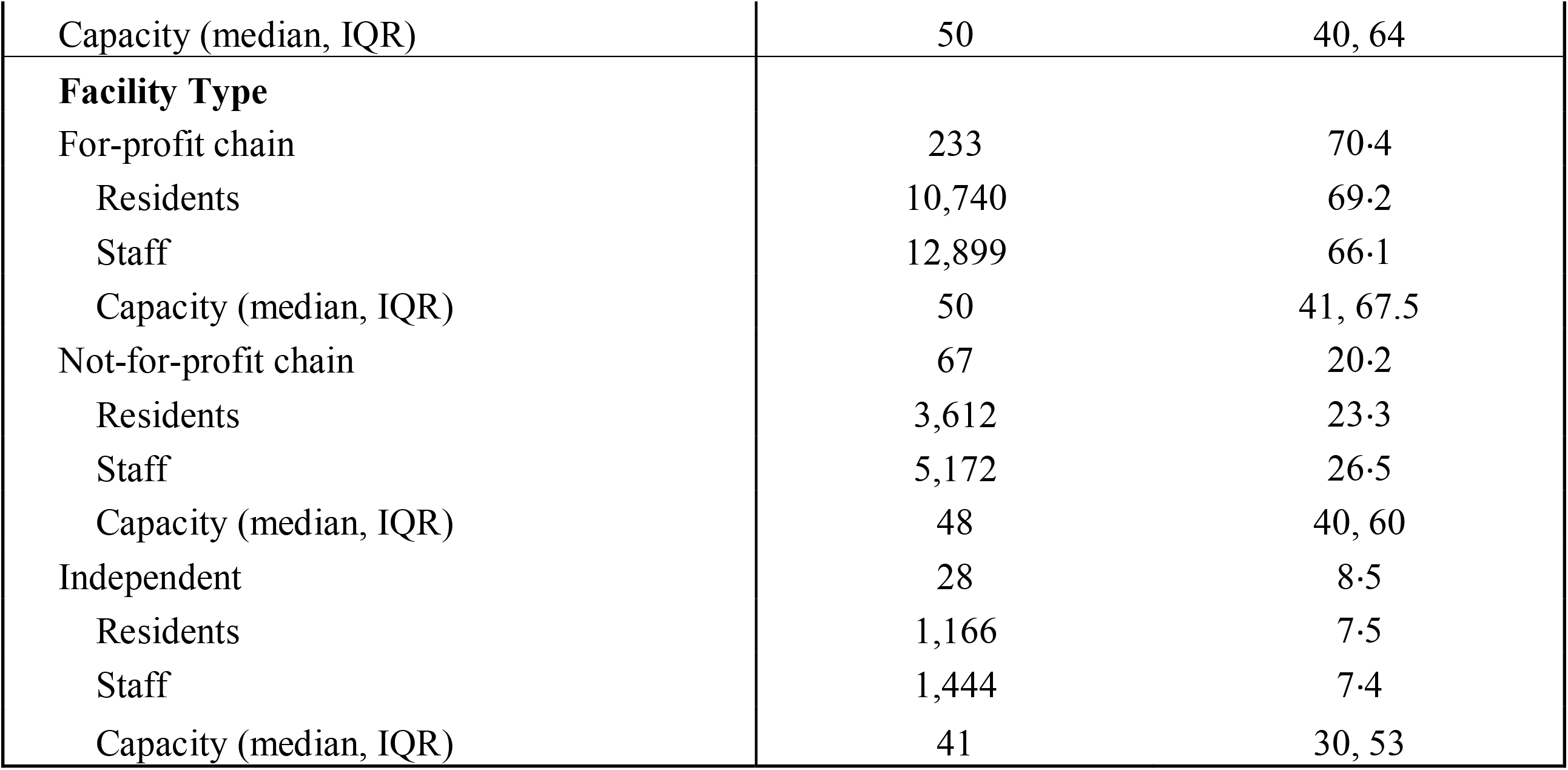
Characteristics of included residents (n=15,518), staff (n=19,515), and LTCFs (n=331)

| **Individuals** | **Residents** |  | **Staff** |  |
| --- | --- | --- | --- | --- |
|  | **n** | **% of total** | **n** | **% of total** |
| **Total** | 15,518 |  | 19,515 |  |
| Age in years (median, IQR) | 87 | 80, 92 | 45 | 32, 56 |
| Male sex | 4,939 | 31.9 | 3,017 | 15.5 |
| Female sex | 10,579 | 68.3 | 16,498 | 84.5 |
| Linked with single LTCF | 15,123 | 97.7 | 18,399 | 94.3 |
| Total follow-up time (median, IQR) | 224 | 59, 349 | 239 | 79, 357 |
| Dose two follow-up time (median, IQR) | 221 | 117, 247 | 173 | 35, 245 |
| **Prior SARS-CoV2 Exposure** |  |  |  |  |
| Pre-analysis PCR positive | 1,180 | 7.6 | 851 | 4.4 |
| Pre-analysis COVID-19 admission | 632 | 4.1 | 70 | 0.4 |
| Pre-vaccination N-antibody results available | 1,421 | 9.2 | 2,758 | 14.1 |
| Pre-vaccination N-antibody positive | 222 | 1.4 | 404 | 2.1 |
| Total with prior infection | 1,780 | 11.5 | 1,281 | 6.6 |
| **PCR testing** |  |  |  |  |
| Total PCR results in at-risk period | 129,965 |  | 373,666 |  |
| PCR results per person per month (mean, SE) | 2.00 | 0.024 | 3.11 | 0.017 |
| Routine PCR tests | 129,682 | 99.79 | 373,660 | 100 |
| Recorded as symptomatic at time of routine test | 487 | 0.38 | 1,475 | 0.39 |
| **Vaccination** |  |  |  |  |
| First dose | 13,302 | 85.7 | 17,348 | 88.9 |
| AstraZeneca (ChAdOx1-S) | 8,844 | 66.5 | 8,048 | 46.4 |
| Pfizer (Comirnaty) | 4,453 | 33.5 | 9,085 | 52.4 |
| Moderna (Spikevax) | 5 | 0.0 | 215 | 1.2 |
| Second dose | 12,082 | 77.9 | 16,612 | 85.1 |
| AstraZeneca (ChAdOx1-S) | 8,052 | 66.6 | 7,620 | 45.9 |
| Pfizer (Comirnaty) | 4,024 | 33.3 | 8,786 | 52.9 |
| Moderna (Spikevax) | 6 | 0.0 | 206 | 1.2 |
| Heterologous primary course | 358 | 2.3 | 146 | 0.7 |
| Primary course vaccine interval | 76 | 68, 79 | 76 | 65, 79 |
| Booster dose | 9,075 | 58.5 | 11,053 | 56.6 |
| AstraZeneca (ChAdOx1-S) | 10 | 0.1 | 9 | 0.1 |
| Pfizer (Comirnaty) | 9,012 | 99.3 | 9,986 | 90.3 |
| Moderna (Spikevax) | 53 | 0.6 | 1,058 | 9.6 |
| Heterologous booster | 5,988 | 38.6 | 5,090 | 26.1 |
| Booster interval | 203 | 193, 217 | 204 | 191, 229 |
| Any heterologous regimen | 6,330 | 40.8 | 5,211 | 26.7 |
| **Long Term Care Facilities** | **n** | **% of total** |  |  |
| **Total** | **331** |  |  |  |

|  |  |  |
| --- | --- | --- |
| Capacity (median, IQR) | 50 | 40, 64 |
| **Facility Type** |  |  |
| For-profit chain | 233 | 70·4 |
| Residents | 10,740 | 69·2 |
| Staff | 12,899 | 66·1 |
| Capacity (median, IQR) | 50 | 41, 67.5 |
| Not-for-profit chain | 67 | 20·2 |
| Residents | 3,612 | 23·3 |
| Staff | 5,172 | 26·5 |
| Capacity (median, IQR) | 48 | 40, 60 |
| Independent | 28 | 8·5 |
| Residents | 1,166 | 7·5 |
| Staff | 1,444 | 7·4 |
| Capacity (median, IQR) | 41 | 30, 53 |

**Figure 1.**
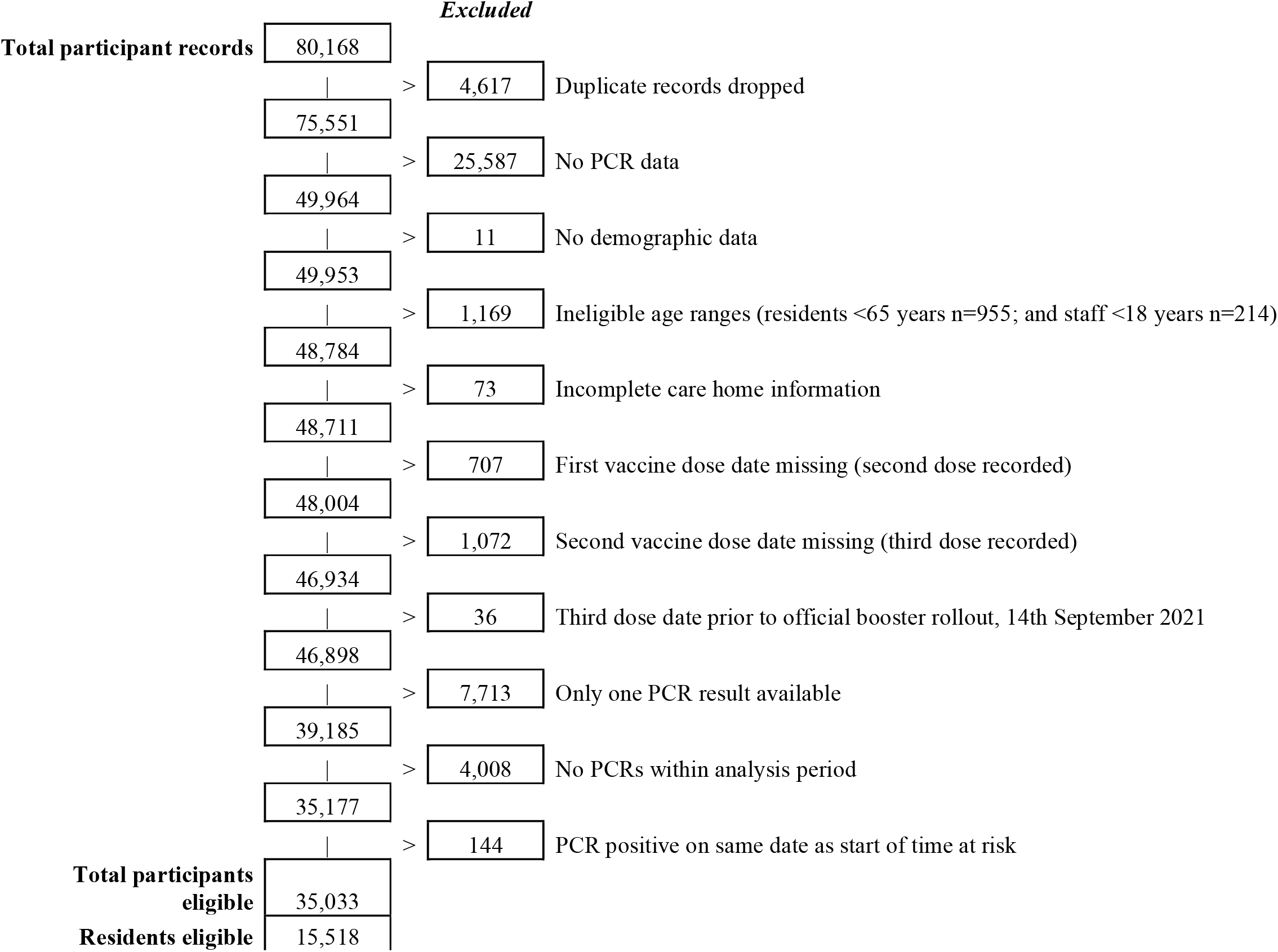
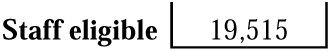
Study inclusion flow diagram

**Figure 2.**
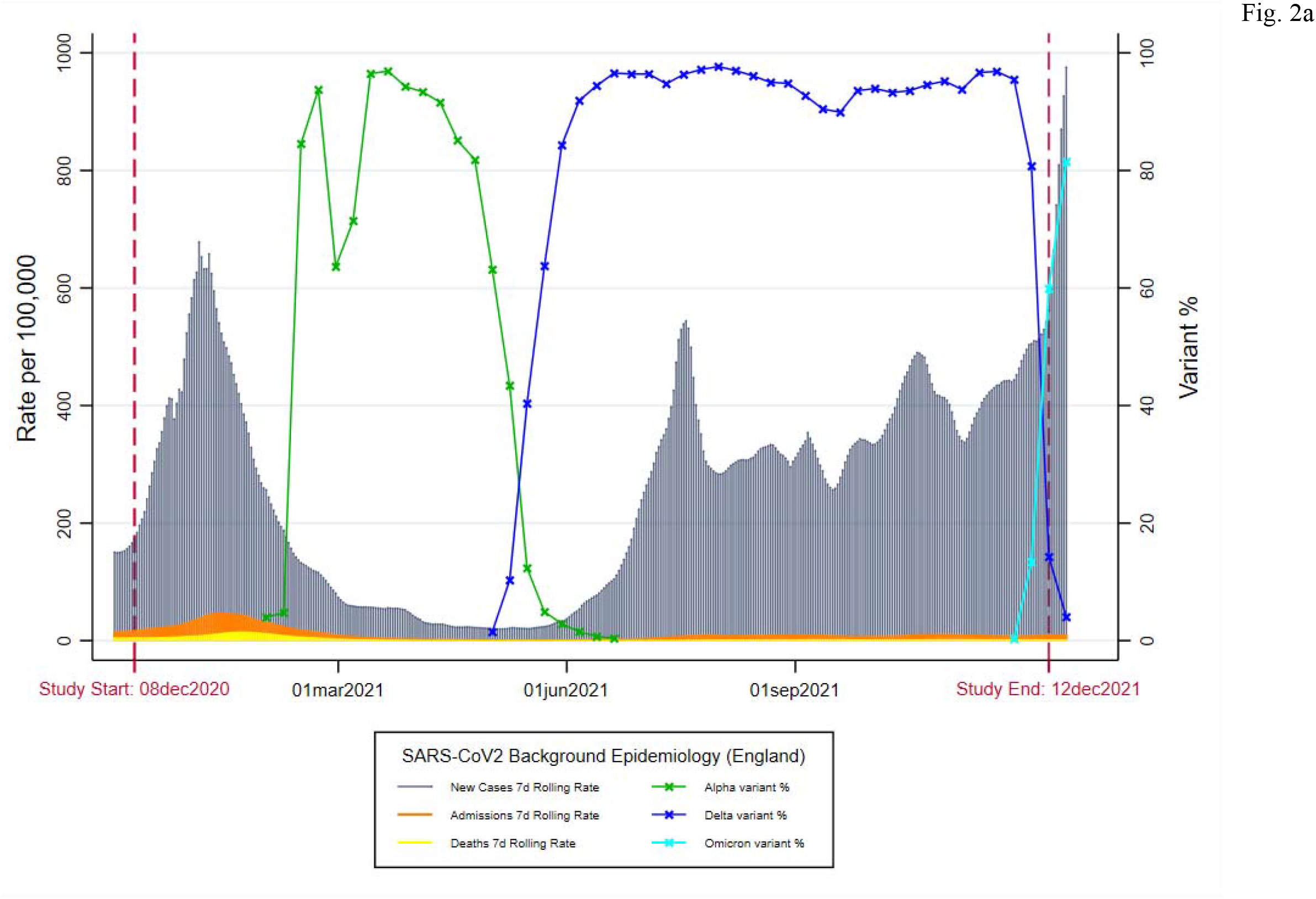
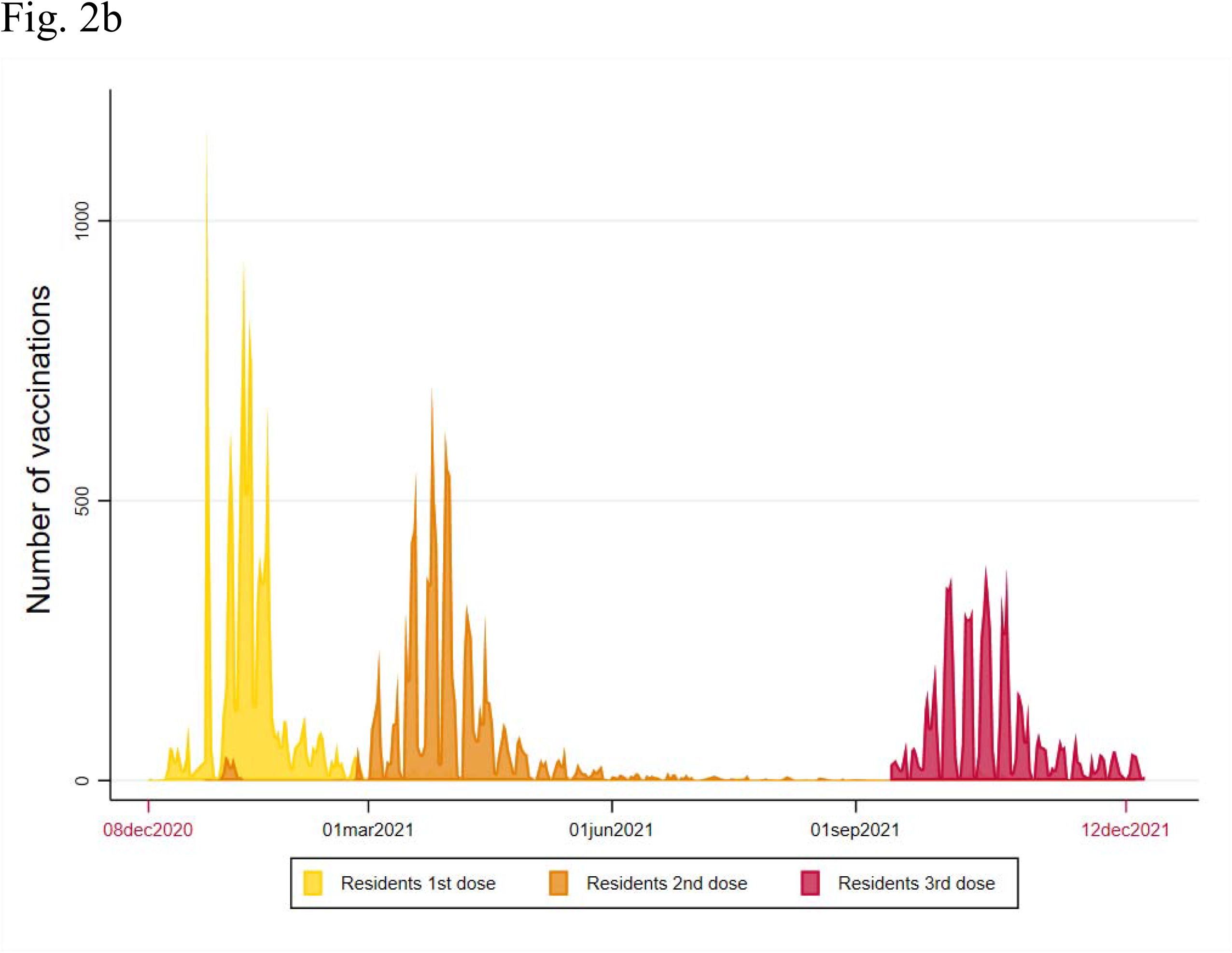
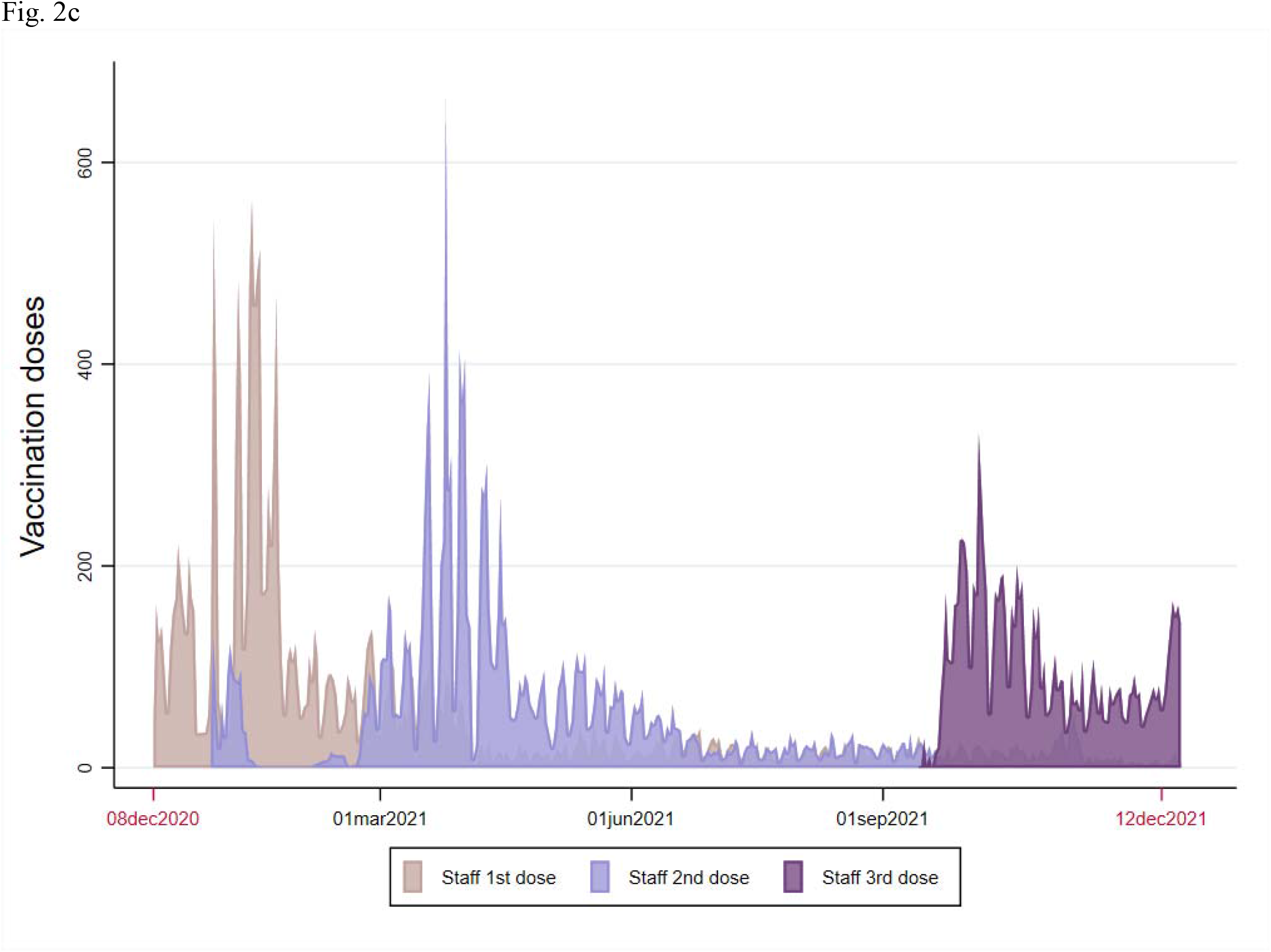
**a)** Background epidemiology of SARS-CoV2 in England by date (Dec 1, 2020 to Dec 16, 2021), including 7-day rolling rates of new cases, hospitalisations, and deaths (data from https://coronavirus.data.gov.uk/); and percentage prevalence over time of the Alpha, Delta, and Omicron variants of concern (data from https://www.gov.uk/government/publications/investigation-of-novel-sars-cov-2-variant-variant-of-concern-20201201). **b)** Numbers of first, second, and third dose vaccinations in included residents by date, over the study period (Dec 8, 2020 to Dec 11, 2021). **c)** Numbers of first, second, and third dose vaccinations in included staff by date, over the study period (Dec 8, 2020 to Dec 11, 2021).

In this cohort, first vaccine doses were mostly administered immediately before Alpha predominance, second doses during Alpha predominance, and booster doses during Delta predominance (Fig 2b, 2c). As their first dose, 66·5% of residents and 46·4% of staff received AZ and 33·5% of residents and 52·4% of staff received Pfizer. The median dose interval was 76 days (IQR 68, 79) for residents and 76 days (65, 79) for staff. Median booster intervals were 203 days (IQR 193, 217) for residents and 204 days (191, 229) for staff.

A mix of for-profit-chains (233/331, 70·4%), not-for-profit chains (67/331, 20·2%), and independent LTCFS (28/331, 8·5%) were included (Table 1). Average capacity varied between 41 beds (30, 53) in independent facilities and 50 (41, 67·5) in for-profit chain facilities.

### Infection

In previously unexposed residents, Cox regression analysis found a significant reduction in hazards of PCR-positive infection at 14-83 days after Dose 2 (VE 50·7%; 95%CI 15·5, 71·3); however, this protective effect was no longer observed at ≥84 days after Dose 2 (Table 2). Protective effects were restored, and optimised, after Dose 3 with VE of 71·6% (53·5, 82·7). This was similar to the protective effect of 80% (65·6, 88·3) seen amongst unvaccinated but previously exposed residents when compared with unvaccinated unexposed residents. In unexposed staff, protective effects of vaccination were seen from 28 days following Dose 1 (25·7% [6·4, 41·1]) with a slight drop seen from 14-83 days (50·3% [32·7, 63·3]) to ≥84 days (42·1% [29·5, 52·4]) after Dose 2 (Table 3). Again, VE peaked after Dose 3 at 78·3% (70·1, 84·3). Significant benefits were seen only after three doses (75·3% [11·6, 93·1]) amongst previously exposed staff. Local area incidence rates were significant predictors of hazards of infection for both residents (aHR 4·462 [2·2, 9·06]) and staff (aHR 5·982 [3·742, 9·563]). Dose 2 VE waning in staff became more apparent when focusing on mRNA recipients (60·7% [44·2, 72·4] at 14-83d vs 45·1% [31·3, 56·2] at ≥84d after Dose 2) (Supplementary Materials). Stratified by primary course type (AZ vs mRNA), VE was similar following Dose 3 for both residents (71·4% [49, 84] vs 71·4 [49·7, 83·8]) and staff (75·9% [61·5, 84·8]) vs 79·3% [70, 85·7]). Censoring at mid-May showed significant VE of 70% (45·3, 83·7) from 28 days after Dose 1 for residents, and 55% (39·5, 66·6) for staff (Supplementary Materials).

**Table 2.**
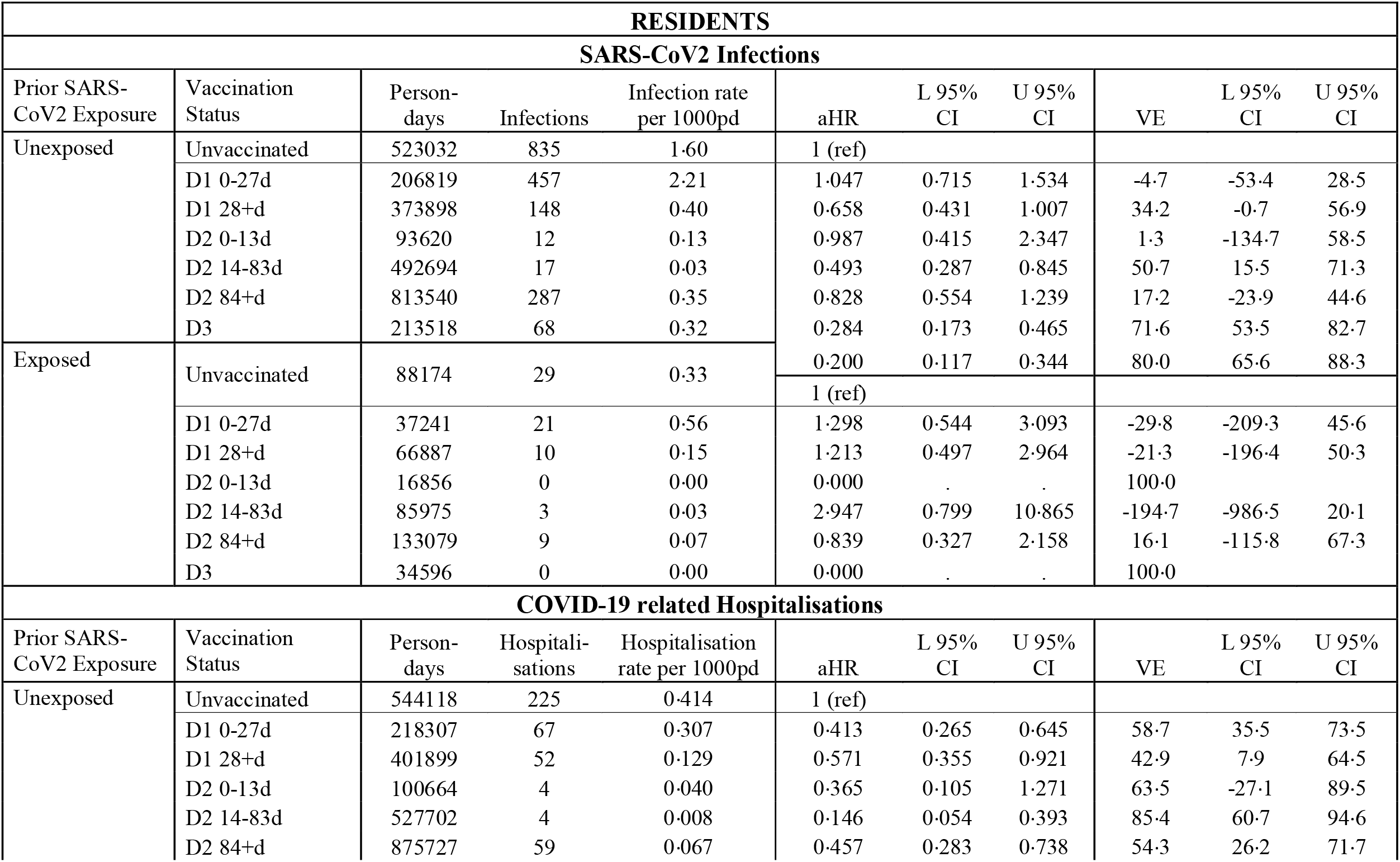
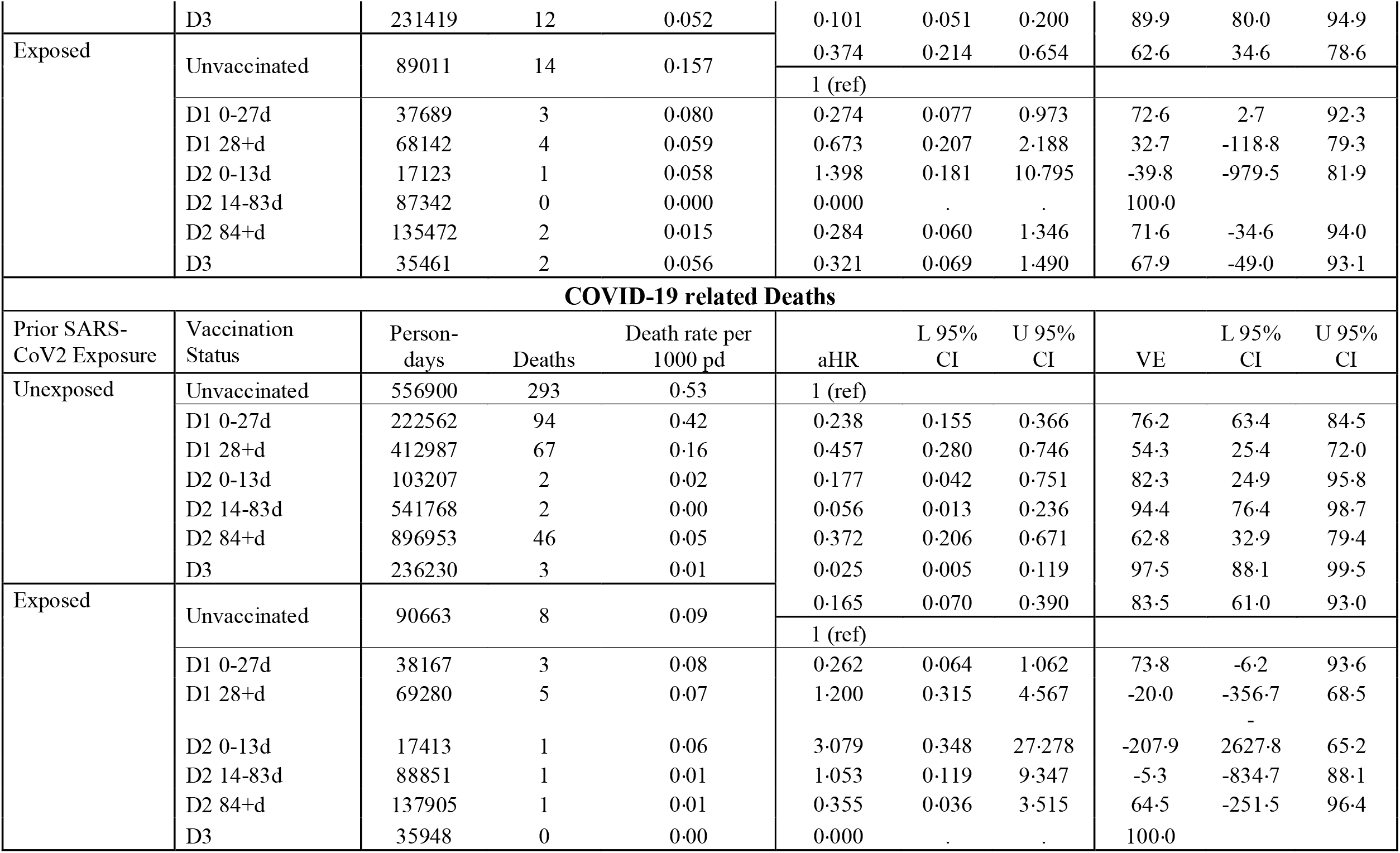
Crude event rates, adjusted hazard ratios, and vaccine effectiveness against PCR-positive SARS-CoV2 Infections, COVID-19 related hospitalisations, and COVID-19 related deaths for LTCF residents, by prior SARS-CoV2 exposure, and vaccination status

**Table 3.**
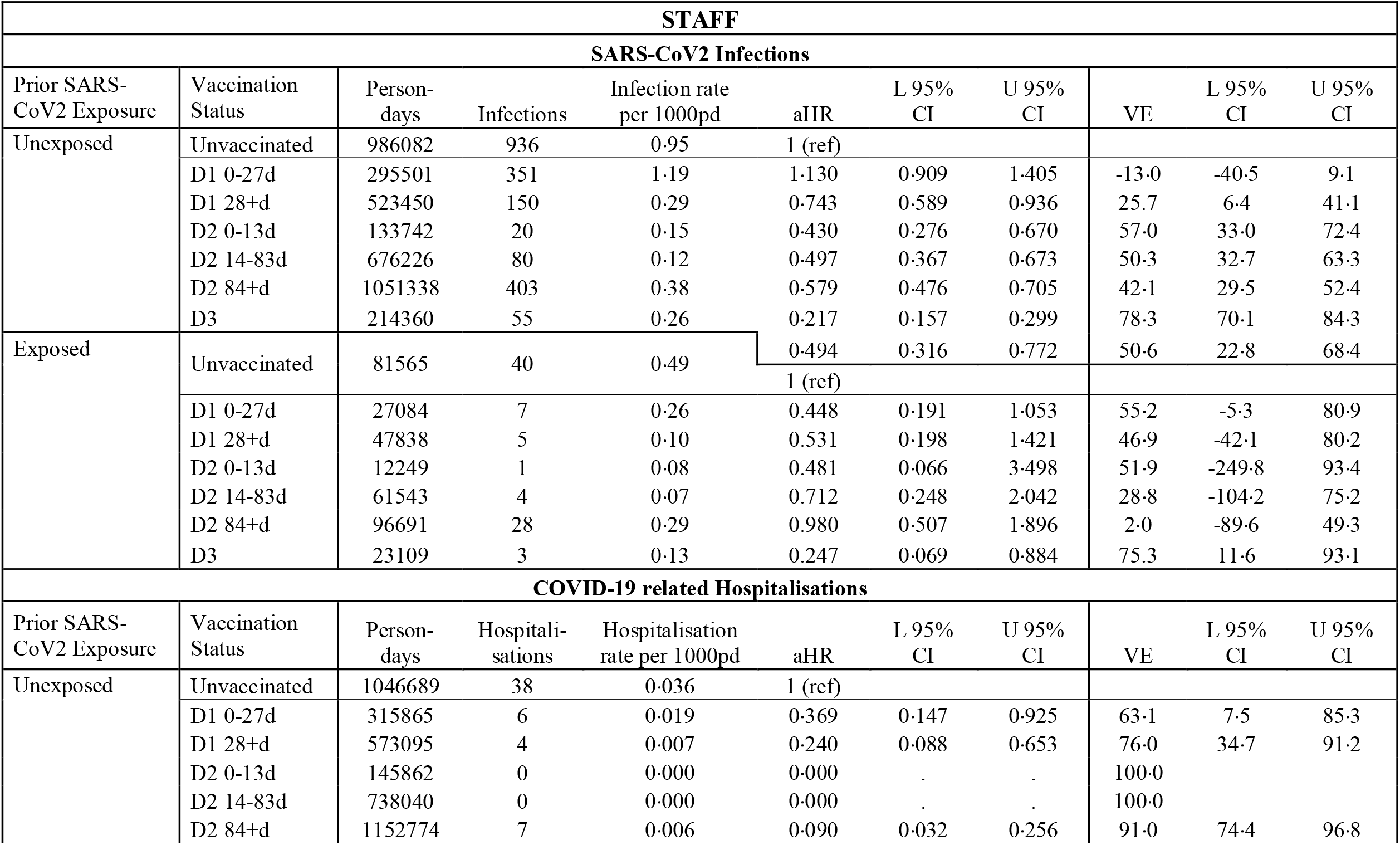
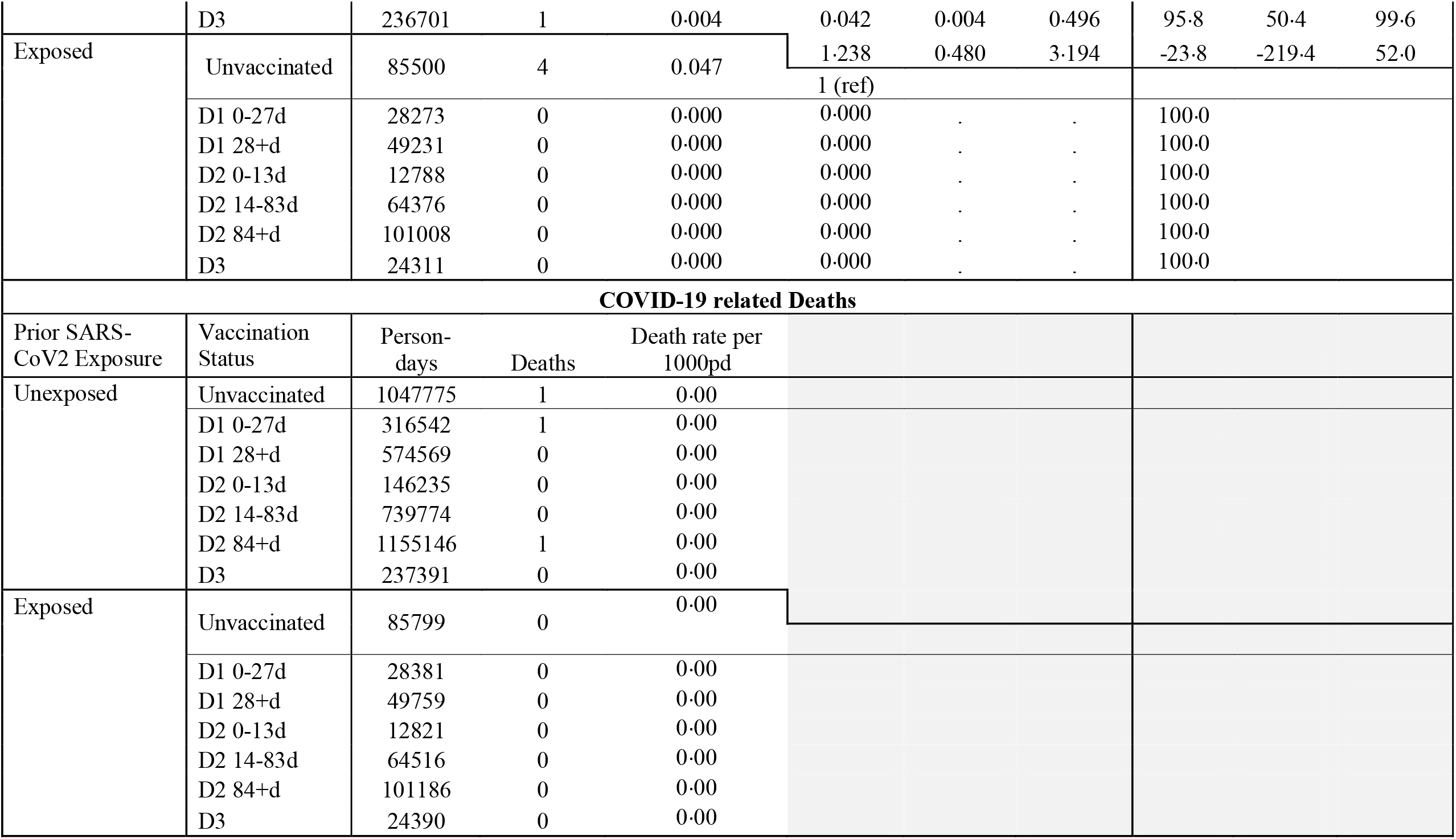
Crude event rates, adjusted hazard ratios, and vaccine effectiveness against PCR-positive SARS-CoV2 Infections, COVID-19 related hospitalisations, and COVID-19 related deaths for LTCF staff, by prior SARS-CoV2 exposure, and vaccination status

### Hospitalisation

There were 449 COVID-19 related hospitalisations in residents, including 225 unvaccinated unexposed and 14 in unvaccinated exposed residents (Table 2). Amongst unexposed residents, vaccination effects were seen immediately after the Dose 1 (58·7% [35·5, 73·5]), rising to 85·4% [60·7, 94·6] at 14-83d after Dose 2 and declining to 54·3% (26·2, 71·7) at ≥84d after Dose 2. VE peaked after three doses in the unexposed (89·9% [80·0, 94·9]), which appeared to be more protective than prior infection in the absence of vaccination (62·6% [34·6, 78·6]). Male residents had higher hazards of hospitalisation than females (aHR 1·97 [1·646, 2·360]), and local area incidence was a strong predictor of hospitalisation in residents (aHR 3·916 [1·597, 9·605]). There were 60 hospitalisations in staff, including 38 in unvaccinated unexposed staff and 4 in unvaccinated exposed staff (Table 3). There appeared to be no substantial waning in immunity against hospitalisations for staff in any of the models, with very few hospitalisations occurring after vaccination. LTCF size was the only covariate with significant effect on hazards of hospitalisation in staff (aHR 0·988 [0·978, 0·997]).

### Death

There were 526 COVID-19 related deaths in residents, including 293 in unvaccinated unexposed residents, and 8 in unvaccinated exposed residents (Table 2). Protective effects in the unexposed were seen immediately after Dose 1 (VE 76·2% [63·4, 84·5]). VE reached 94·4% (76·4, 98·7) at 14-83d after Dose 2, but declined from ≥84d to 62·8% (32·9, 79·4). The third dose boosted VE to 97·5% (88·1, 99·5). Male sex (aHR 1·988 [1·648, 2·399]), LTCF size (aHR 0·994 [0·998, 0·999]), and local area incidence (aHR 6·188 [2·761, 13·866]) were all significantly associated with hazards of death. Due to small event numbers it was not possible to model VE against deaths for staff (Table 3).

### Ct Values

The mean Ct values from unvaccinated individuals (Nucleocapsid: 25·92 [SD 7·32]) were lower than those at ≥28 days after Dose 1 (29·44 [SD 9·8]; p<0·0001) and 14-83 days after Dose 2 (30·86 [SD 7·07], p<0·0001), when Alpha was predominant. However, Ct values from later time periods when Delta was predominant, were not significantly different from unvaccinated samples (Supplementary Materials). Ct values amongst unvaccinated individuals were significantly lower during Delta predominance than pre-Delta (Nucleocapsid: 26·28 [sd 7·37] vs 23·58 [6·59]; p=0·0021) (Supplementary Materials).

## Discussion

In this prospective cohort study of 15,518 older residents and 19,515 staff from 331 LTCFs across England, we found that the moderate protection against infection and the high protection against hospitalisation and death in the 2-12 weeks following completion of a primary course of AstraZeneca or mRNA vaccination declines markedly beyond 12 weeks. This is seen for all three outcomes in residents and also against infection amongst staff. High levels of protection were restored, and maximised, following a third ‘booster’ dose, with VE of 71·6% and 78·3% against infection, and 89·9% and 95·8% against hospitalisation, for residents and staff respectively, and 97·5% against death for residents. Though this analysis took place in the pre-Omicron period, these trends of waning immunity over time are likely to be generalisable across variants, carrying important implications for long-term vaccination policy in LTCFs.

Our estimates for short-term Dose 1 and 2 VE against infection and severe clinical outcomes are broadly similar to other studies in LTCF residents and staff,^6-12^ however the majority of these were prior to emergence of the Delta variant, and had insufficient follow-up time to examine durability of two-dose immunity. We were able to follow-up staff and residents for 24-32 weeks following Dose 2, which is substantially longer than previous studies, making it possible to demonstrate waning in two-dose VE after 12 weeks and the restorative impact of boosters. VE in staff did not decline in a similar fashion to residents, with more modest decreases in VE against infection, and none against hospitalisation, in line with other population-based studies that reported waning of immunity against infection but not against severe outcomes.^24–26^ In contrast to these studies, and to results from our staff cohort, we found that VE against severe clinical outcomes did wane substantially in residents, indicating a significant underlying difference between LTCF residents and the wider population.

Waning of VE against severe outcomes in residents may be driven by immunosenescence and their clinical frailty, an explanation which is supported by data suggesting greater waning in older or more clinically vulnerable groups.^18^ Differences between staff and residents may also be partly driven by vaccine type, as staff were vaccinated predominantly with mRNA vaccines while residents with AstraZeneca, however U.K. studies vaccines have generally shown significant waning following both AstraZeneca and Pfizer vaccines, though with Pfizer generally starting from higher peak VE.^16,18^ These findings of waning after Pfizer are also consistent with VE studies from Israel^17^, and with immunological data from the U.K.^13^ Whatever the underlying drivers of these novel findings amongst residents, the observation that vaccine-derived immunity against hospitalisation and death wanes over time constitutes significant cause for concern.

Our findings are concordant with emerging clinical effectiveness data^26^ and immunological data^27,28^ demonstrating the additional benefit of a booster dose, and with data from Israel showing reduced infection rates in LTCF residents in the time period following roll-out of boosters.^29^ Furthermore, three vaccine doses in unexposed residents appeared to offer equal or higher protection compared with prior exposure in the absence of vaccination, and we found rates of all three outcomes to be lowest in those with both prior exposure and vaccination. We were also able to compare VE for previously unexposed residents and staff who received AZ vs mRNA (largely Pfizer) primary courses followed by a third mRNA dose, corresponding to two natural cohorts of heterologous and homologous booster recipients, respectively. We found booster VE to be roughly equal between the two cohorts, with a suggestion of slightly higher VE against hospitalisation for the heterologous resident group (AZ primary course + mRNA booster). Together, these findings highlight the importance of a third booster dose irrespective of prior exposure status or primary course type.

In contrast to an earlier analysis, where we estimated VE against infection to be 56% at 4-5 weeks, and 62% at 5-6 weeks following Dose 1 in residents,^30^ we did not observe any significant first-dose VE in the primary analysis; however after excluding censoring the analysis in May, VE estimates against infection at ≥28d after Dose 1 became more consistent with our previous results. This is in keeping with the reduced vaccine efficacy against Delta variant infections that has been reported elsewhere.^16^

Strengths of this study were the use of high-quality routinely collected national data on testing, vaccination, hospitalisations, and death registrations for a large cohort of staff and residents. This population underwent frequent asymptomatic PCR testing through their LTCFs, which allowed us to reliably identify eligible residents and staff and accurately define their time-at-risk.

Limitations included the inability to distinguish hospitalisations and deaths that occurred ‘with’ rather than ‘from’ COVID-19, particularly for frail and comorbid residents, which is likely to underestimate VE; the probable significant under-ascertainment of prior exposure, particularly amongst staff, which is also likely to underestimate VE; and the possible under-ascertainment of infection events detected on LFDs without PCR confirmation, particularly amongst staff, which may overestimate VE against infection.

Based on these data, and what is already known about immunity waning, we expect the reported trends to be generalisable over future rounds of vaccinations and variants. The available evidence suggests that while vaccines can induce powerful short-term immunity, this protection may quickly fade. While the risk of severe outcomes currently appears lower for the Omicron variant,^31, 32^ this risk is likely to increase with waning immunity, particularly for the frail LTCF resident cohort. Our data suggest that regular boosting may be required to maintain protective effects over a longer term; however, given the complex economic and political backdrop to vaccine procurement and distribution, it will be important to consider how vulnerable groups can continue to be protected against COVID-19 whilst ensuring global vaccine equity. Our findings also highlight the critical importance of ongoing surveillance in LTCF residents, to provide early warning of surges in infection associated with waning immunity and the emergence of further new variants.

## Data Availability

De-identified test results and limited metadata will be made available for use by researchers in future studies, subject to appropriate research ethical approvals once the VIVALDI study cohort has been finalised. These datasets will be accessible via the Health Data Research UK Gateway.

## Contributors

MS, LS, AC, and MK conceptualised the study. MS, AC, LS and MK developed the statistical analysis plan. MS did the formal statistical analysis. MK, CF, BA, HNL, VB and AI-S were involved with project administration. LS and AH obtained research funding. MS wrote the first draft of the manuscript. All authors revised and edited the manuscript. AC, MS, LS and MK accessed and verified the data. All authors had full access to all the data reported in the study. LS, AC, and MS shared the final responsibility for the decision to submit for publication.

## Acknowledgements

We thank the staff and residents in the LTCFs that participated in this study and Mark Marshall at NHS England who pseudonymised the electronic health records.

## References

1 Morciano M, Stokes J, Kontopantelis E, Hall I, Turner AJ. Excess mortality for care home residents during the first 23 weeks of the COVID-19 pandemic in England: a national cohort study. BMC Med 2021; 19: 1–11.

2 Castro-Herrera VM, Lown M, Fisk HL, et al. Relationships Between Age, Frailty, Length of Care Home Residence and Biomarkers of Immunity and Inflammation in Older Care Home Residents in the United Kingdom. Front Aging 2021; 2. DOI:10.3389/fragi.2021.599084.

3 Independent report - Priority groups for coronavirus (COVID-19) vaccination: advice from the JCVI. 30 December 2020. https://www.gov.uk/government/publications/priority-groups-for-coronavirus-covid-19-vaccination-advice-from-the-jcvi-30-december-2020. Accessed 23 February 2022.

4 Independent report - JCVI statement regarding a COVID-19 booster vaccine programme for winter 2021 to 2022 coronavirus. 14 September 2021 https://www.gov.uk/government/publications/jcvi-statement-september-2021-covid-19-booster-vaccine-programme-for-winter-2021-to-2022/. Accessed 23 February 2022.

5 Helfand BKI, Webb M, Gartaganis SL, Fuller L, Kwon C-S, Inouye SK. The Exclusion of Older Persons From Vaccine and Treatment Trials for Coronavirus Disease 2019—Missing the Target. JAMA Intern Med 2020; 180: 1546.

6 Subbarao AS, Copas A, Andrews N, et al. Vaccine Effectiveness Against Infection and Death Due to SARS-CoV-2, Following One and Two Doses of the BNT162b2 and ChADox-1 in Residents of Long-Term Care Facilities in England, Using a Time-Varying Proportional Hazards Model. SSRN Electron J 2021.

7 Mazagatos C, Monge S, Olmedo C, et al. Effectiveness of mRNA COVID-19 vaccines in preventing SARS-CoV-2 infections and COVID-19 hospitalisations and deaths in elderly long-term care facility residents, Spain, weeks 53 2020 to 13 2021. Eurosurveillance 2021; 26: 1–6.

8 Monge S, Olmedo C, Alejos B, Lapeña MF, Sierra MJ, Limia; A. Direct and Indirect Eff ectiveness of mRNA Vaccination against Severe Acute Respiratory Syndrome Coronavirus 2 in Long-Term Care Facilities, Spain. Emerg Infect Dis; 2021.

9 Lefèvre B, Tondeur L, Madec Y, et al. Beta SARS-CoV-2 variant and BNT162b2 vaccine effectiveness in long-term care facilities in France. Lancet Heal Longev 2021; 2: e685–7.

10 Nanduri S, Pilishvili T, Derado G, et al. Effectiveness of Pfizer-BioNTech and Moderna Vaccines in Preventing SARS-CoV-2 Infection Among Nursing Home Residents Before and During Widespread Circulation of the SARS-CoV-2 B.1.617.2 (Delta) Variant - National Healthcare Safety Network, March 1-August. MMWR Morb Mortal Wkly Rep 2021; 70: 1163–6.

11 Starrfelt J, Danielsen AS, Kacelnik O, Børseth AW, Seppälä E, Meijerink H. High vaccine effectiveness against COVID-19 infection and severe disease among residents and staff of long-term care facilities in Norway, November – June 2021. medRxiv 2021. https://doi.org/10.1101/2021.08.08.21261357.

12 Moustsen-Helms IR, Emborg H-D, Nielsen J, et al. Vaccine effectiveness after 1st and 2nd dose of the BNT162b2 mRNA Covid-19 Vaccine in long-term care facility residents and healthcare workers – a Danish cohort study. medRxiv 2021.

13 Aldridge RW, Yavlinsky A, Nguyen VG, et al. Waning of SARS-CoV-2 antibodies targeting the Spike protein in individuals post second dose of ChAdOx1 and BNT162b2 COVID-19 vaccines and risk of breakthrough infections: analysis of the Virus Watch community cohort. medRxiv 2021; 2021.11.05.21265968.

14 Levin EG, Lustig Y, Cohen C, et al. Waning Immune Humoral Response to BNT162b2 Covid-19 Vaccine over 6 Months. N Engl J Med 2021; 385: e84.

15 Wei J, Pouwels KB, Stoesser N, et al. SARS-CoV-2 anti-spike IgG antibody responses after second dose of ChAdOx1 or BNT162b2 and correlates of protection in the UK general population. medRxiv 2021; : 2021.09.13.21263487.

16 Pouwels KB, Pritchard E, Matthews PC, et al. Effect of Delta variant on viral burden and vaccine effectiveness against new SARS-CoV-2 infections in the UK. Nat Med 2021; 27: 2127–35.

17 Goldberg Y, Mandel M, Bar-On YM, et al. Waning Immunity after the BNT162b2 Vaccine in Israel. N Engl J Med 2021; 385: e85.

18 Andrews N, Tessier E, Stowe J, et al. Duration of Protection against Mild and Severe Disease by Covid-19 Vaccines. N Engl J Med 2022; 386: 340–50.

19 UK Health Security Agency. Investigation of Novel SARS-CoV-2 Variants of Concern. https://www.gov.uk/government/publications/investigation-of-novel-sars-cov-2-variant-variant-of-concern-20201201.

20 Krutikov M, Palmer T, Donaldson A, et al. Study Protocol: Understanding SARS-Cov-2 infection, immunity and its duration in care home residents and staff in England (VIVALDI). Wellcome Open Res 2021; 5: 232.

21 NHS COVID-19 Data Store. https://data.england.nhs.uk/covid-19/. Accessed 23 February 2022.

22 UK Health Security Agency. Coronavirus (COVID-19) in the UK dashboard. https://coronavirus.data.gov.uk/. Accessed 23 February 2022.

23 UK Department of Health and Social Care. Vivaldi study: privacy notice. 24 March 2021. https://www.gov.uk/government/publications/vivaldi-study-privacy-notice/vivaldi-study-privacy-notice. Accessed 23 February 2022.

24 Chemaitelly H, Tang P, Hasan MR, et al. Waning of BNT162b2 Vaccine Protection against SARS-CoV-2 Infection in Qatar. N Engl J Med 2021; 385: e83.

25 Tartof SY, Slezak JM, Fischer H, et al. Effectiveness of mRNA BNT162b2 COVID-19 vaccine up to 6 months in a large integrated health system in the USA: a retrospective cohort study. Lancet 2021; 398: 1407–16.

26 Bar-On YM, Goldberg Y, Mandel M, et al. Protection against Covid-19 by BNT162b2 Booster across Age Groups. N Engl J Med 2021; 385: 2421–30.

27 Yavlinsky A, Beale S, Nguyen V, et al. Anti-spike antibody trajectories in individuals previously immunised with BNT162b2 or ChAdOx1 following a BNT162b2 booster dose. medRxiv 2022.

28 Tut G, Lancaster T, Krutikov M, et al. Booster Vaccination Strongly Enhances SARS-CoV-2-Specific Antibody and Cellular Responses in Elderly Residents of Care Homes. SSRN Electron J 2021.

29 Muhsen K, Maimon N, Mizrahi A, et al. Effects of BNT162b2 Covid-19 Vaccine Booster in Long-Term Care Facilities in Israel. N Engl J Med 2022; 386: 399–401.

30 Shrotri M, Krutikov M, Palmer T, et al. Vaccine effectiveness of the first dose of ChAdOx1 nCoV-19 and BNT162b2 against SARS-CoV-2 infection in residents of long-term care facilities in England (VIVALDI): a prospective cohort study. Lancet Infect Dis 2021; 21: 1529–38.

31 Wolter N, Jassat W, Walaza S, et al. Early assessment of the clinical severity of the SARS-CoV-2 omicron variant in South Africa: a data linkage study. The Lancet 2022.

32 Krutikov M, Stirrup O, Nacer-Laidi H, et al. Outcomes of SARS-CoV-2 Omicron infection in residents of Long-Term Care. medRxiv 2022.

