## Supplementary appendix for "Duration of vaccine effectiveness against SARS-CoV2 infection, hospitalisation, and death in residents and staff of Long-Term Care Facilities (VIVALDI): a prospective cohort study, England, Dec 2020-Dec 2021"

### SUPPLEMENTARY MATERIAL

**Figure S1**

Cycle threshold values of a) Nucleocapsid and b) ORF1ab gene targets from viral isolates, by vaccination status

**Figure S2**

Ct values of a) Nucleocapsid and b) ORF1ab gene targets in unvaccinated individuals before and during Delta variant predominance.

**Table S1**

Crude rates, adjusted hazard ratios, and vaccine effectiveness against infections, stratified by vaccine type, in unexposed residents and staff.

**Table S2**

Crude rates, adjusted hazard ratios, and vaccine effectiveness against infections in the pre-Delta period in unexposed residents and staff.

**Table S3**

Cycle threshold values of Nucleocapsid and ORF1ab gene targets from viral isolates, by vaccination status across the whole study period; and in unvaccinated individuals before and during Delta variant predominance.

**Table S4**

Crude rates, adjusted hazard ratios, and vaccine effectiveness against hospitalisations, stratified by vaccine type, in unexposed residents and staff.

**Table S5**

Crude rates, adjusted hazard ratios, and vaccine effectiveness against deaths, stratified by vaccine type, in unexposed residents.

**Fig S1a**

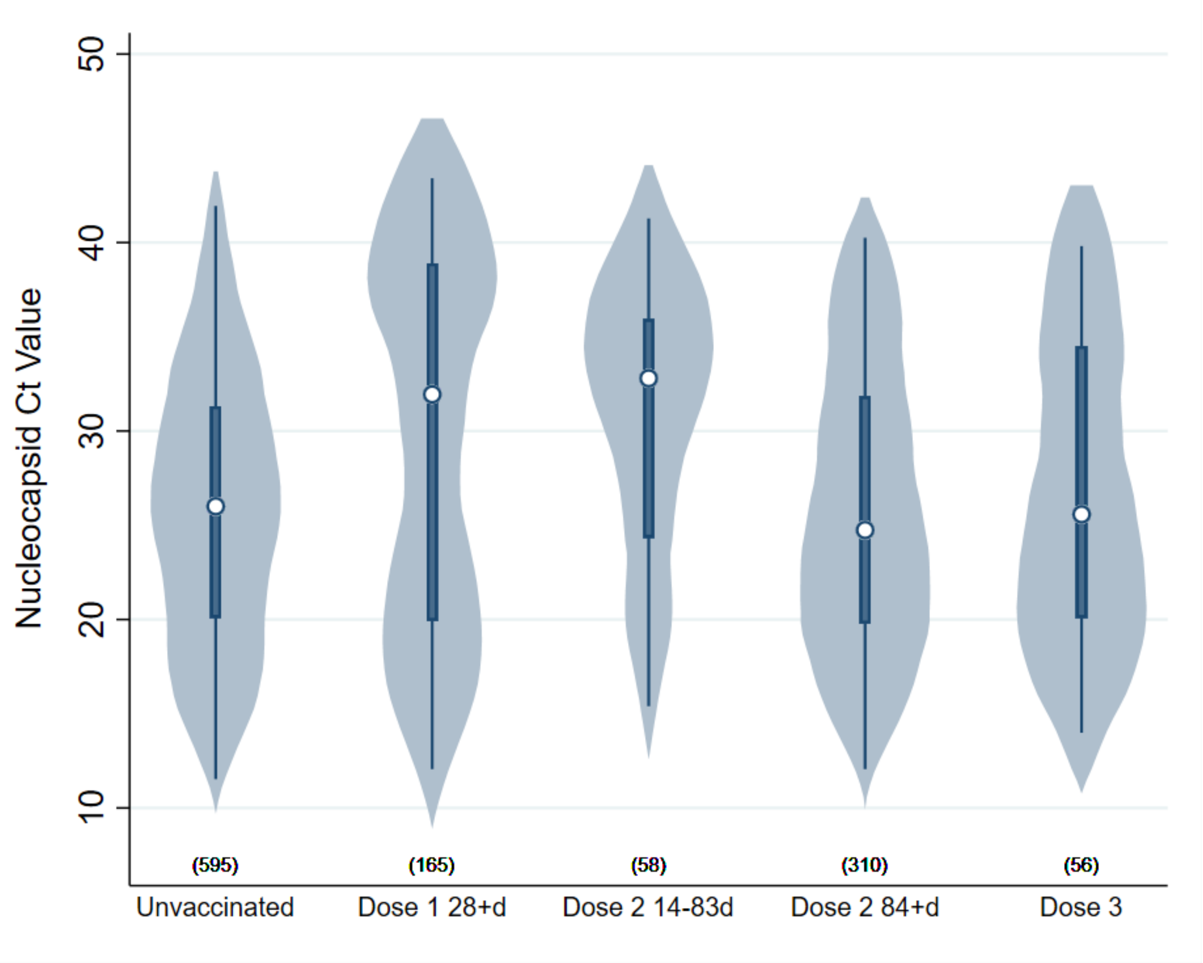

Nucleocapsid Ct Value
50
40
30
20
10
(595)
(165)
(58)
(310)
(56)
Unvaccinated
Dose 1 28+d
Dose 2 14-83d
Dose 2 84+d
Dose 3

**Fig S1b**

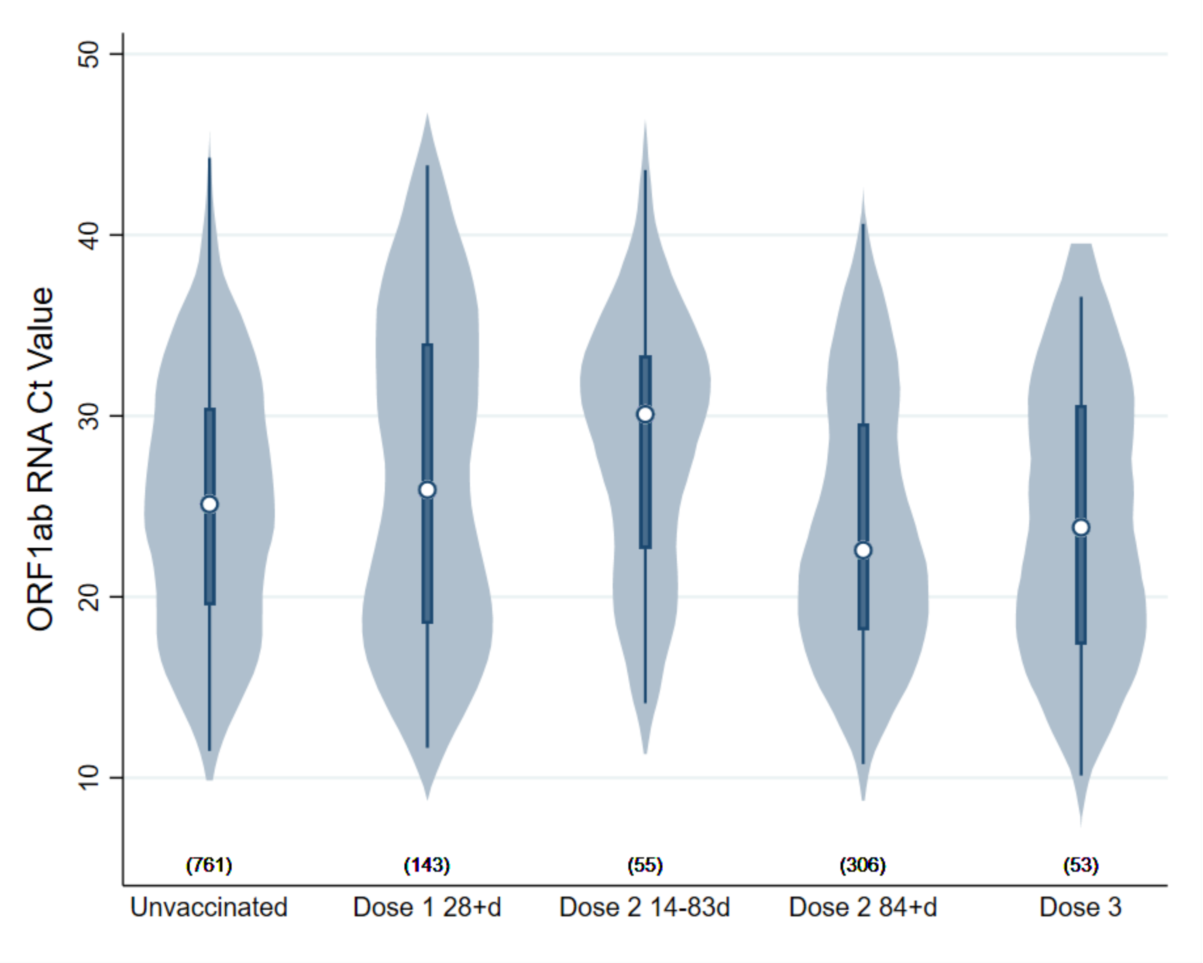
ORF1ab RNA Ct Value
50
40
30
20
10
(761)
(143)
(55)
(306)
(53)
Unvaccinated
Dose 1 28+d
Dose 2 14-83d
Dose 2 84+d
Dose 3

**Figure S2a**

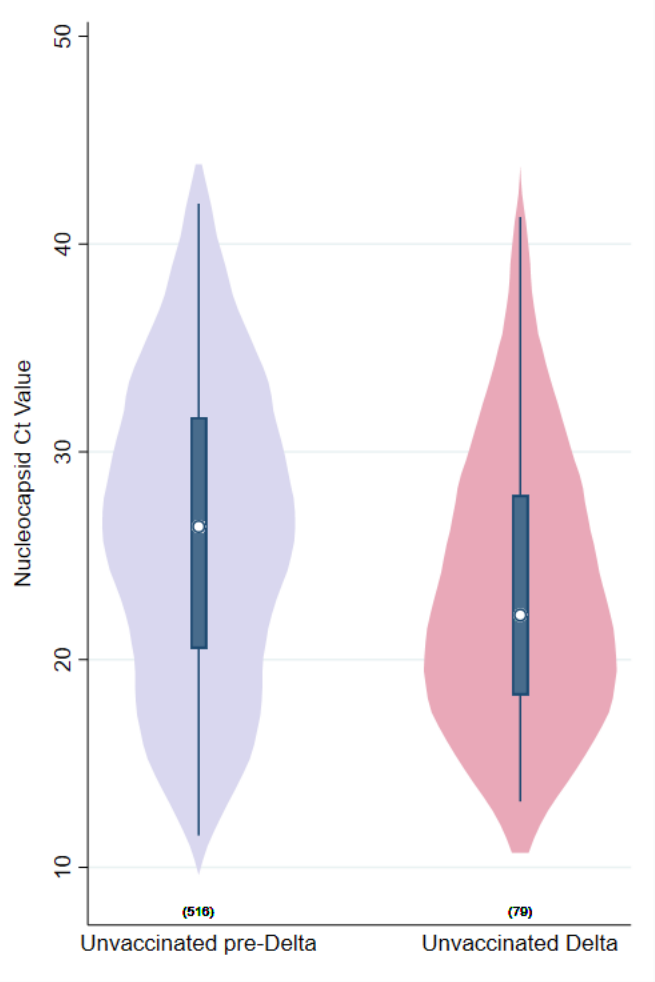
Nucleocapsid Ct Value
50
40
30
20
10
(516)
(79)
Unvaccinated pre-Delta
Unvaccinated Delta

**Figure S2b**

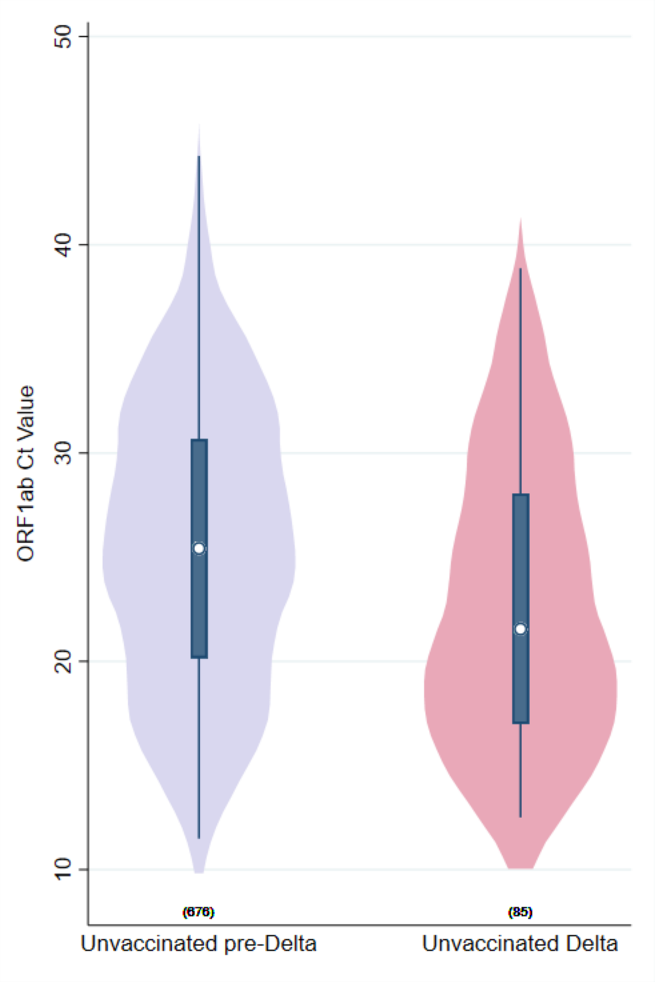
ORF1ab Ct Value
50
40
30
20
10
(676)
(85)
Unvaccinated pre-Delta
Unvaccinated Delta

**Table S1**

| Prior SARS-CoV2 Status | Vaccination Status | Person-days | Infection Events | Infection rate per 1000 person-days | aHR | L 95% CI | U 95% CI | VE | L 95% CI | U 95% CI |
|---|---|---|---|---|---|---|---|---|---|---|
| **Residents** | | | | | | | | | | |
| Unexposed | Unvaccinated | 523032 | 835 | 1.60 | 1 (ref) | | | | | |
| | AZ D1 0-27d | 141935 | 263 | 1.85 | 0.938 | 0.617 | 1.426 | 6.2 | -42.6 | 38.3 |
| | AZ D1 28+d | 261294 | 81 | 0.31 | 0.587 | 0.378 | 0.912 | 41.3 | 8.8 | 62.2 |
| | AZ D2 0-13d | 64982 | 11 | 0.17 | 1.978 | 0.713 | 5.484 | -97.8 | -448.4 | 28.7 |
| | AZ D2 14-83d | 341448 | 9 | 0.03 | 0.379 | 0.164 | 0.879 | 62.1 | 12.1 | 83.6 |
| | AZ D2 84+d | 539330 | 207 | 0.38 | 0.864 | 0.561 | 1.332 | 13.6 | -33.2 | 43.9 |
| | AZ D3 | 132964 | 43 | 0.32 | 0.286 | 0.160 | 0.510 | 71.4 | 49.0 | 84.0 |
| | mRNA D1 0-27d | 64884 | 194 | 2.99 | 1.164 | 0.707 | 1.917 | -16.4 | -91.7 | 29.3 |
| | mRNA D1 28+d | 112604 | 67 | 0.60 | 0.683 | 0.409 | 1.142 | 31.7 | -14.2 | 59.1 |
| | mRNA D2 0-13d | 28638 | 1 | 0.03 | 0.157 | 0.022 | 1.111 | 84.3 | -11.1 | 97.8 |
| | mRNA D2 14-83d | 151246 | 8 | 0.05 | 0.745 | 0.353 | 1.575 | 25.5 | -57.5 | 64.7 |
| | mRNA D2 84+d | 274210 | 80 | 0.29 | 0.737 | 0.446 | 1.217 | 26.3 | -21.7 | 55.4 |
| | mRNA D3 | 80554 | 25 | 0.31 | 0.286 | 0.162 | 0.503 | 71.4 | 49.7 | 83.8 |

| Prior SARS-CoV2 Status | Vaccination Status | Person-days | Infection Events | Infection rate per 1000 person-days | aHR | L 95% CI | U 95% CI | VE | L 95% CI | U 95% CI |
|---|---|---|---|---|---|---|---|---|---|---|
| **Staff** | | | | | | | | | | |
| Unexposed | Unvaccinated | 986082 | 936 | 0.95 | 1 (ref) | | | | | |
| | AZ D1 0-27d | 134378 | 134 | 1.00 | 1.253 | 0.909 | 1.728 | -25.3 | -72.8 | 9.1 |
| | AZ D1 28+d | 267114 | 56 | 0.21 | 0.804 | 0.584 | 1.107 | 19.6 | -10.7 | 41.6 |
| | AZ D2 0-13d | 60405 | 3 | 0.05 | 0.524 | 0.168 | 1.637 | 47.6 | -63.7 | 83.2 |
| | AZ D2 14-83d | 313395 | 41 | 0.13 | 0.710 | 0.457 | 1.103 | 29.0 | -10.3 | 54.3 |
| | AZ D2 84+d | 464836 | 217 | 0.47 | 0.631 | 0.501 | 0.794 | 36.9 | 20.6 | 49.9 |

| | | | | | | | | | | |
|---|---|---|---|---|---|---|---|---|---|---|
| | AZ D3 | 73145 | 21 | 0.29 | 0.241 | 0.152 | 0.385 | 75.9 | 61.5 | 84.8 |
| | mRNA D1 0-27d | 161123 | 217 | 1.35 | 1.072 | 0.831 | 1.384 | -7.2 | -38.4 | 16.9 |
| | mRNA D1 28+d | 256336 | 94 | 0.37 | 0.730 | 0.557 | 0.956 | 27.0 | 4.4 | 44.3 |
| | mRNA D2 0-13d | 73337 | 17 | 0.23 | 0.420 | 0.258 | 0.681 | 58.0 | 31.9 | 74.2 |
| | mRNA D2 14-83d | 362831 | 39 | 0.11 | 0.393 | 0.276 | 0.558 | 60.7 | 44.2 | 72.4 |
| | mRNA D2 84+d | 586502 | 186 | 0.32 | 0.549 | 0.438 | 0.687 | 45.1 | 31.3 | 56.2 |
| | mRNA D3 | 141215 | 34 | 0.24 | 0.207 | 0.143 | 0.300 | 79.3 | 70.0 | 85.7 |

**Table S2**

| Residents | | | | | | | | | | |
|---|---|---|---|---|---|---|---|---|---|---|
| Prior SARS-CoV2 Status | Vaccination Status | Person-days | Infection Events | Infection rate per 1000 person-days | aHR | L 95% CI | U 95% CI | VE | L 95% CI | U 95% CI |
| Unexposed | Unvaccinated | 315640 | 788 | 2.5 | 1 (ref) | | | | | |
| | D1 0-27d | 203526 | 455 | 2.24 | 0.729 | 0.468 | 1.136 | 27.1 | -13.6 | 53.2 |
| | D1 28+d | 315271 | 132 | 0.42 | 0.300 | 0.163 | 0.547 | 70.0 | 45.3 | 83.7 |
| | D2 0-13d | 87389 | 11 | 0.13 | 0.406 | 0.133 | 1.241 | 59.4 | -24.1 | 86.7 |
| | D2 14-83d | 17084 | 5 | 0.29 | 0.760 | 0.334 | 1.732 | 24.0 | -73.2 | 66.6 |
| | D2 84+d | 62 | 0 | 0 | 0.000 | . | . | 100.0 | | |
| **Staff** | | | | | | | | | | |
| Prior SARS-CoV2 Status | Vaccination Status | Person-days | Infection Events | Infection rate per 1000 person-days | aHR | L 95% CI | U 95% CI | VE | L 95% CI | U 95% CI |
| Unexposed | Unvaccinated | 513181 | 840 | 1.64 | 1 (ref) | | | | | |
| | D1 0-27d | 261759 | 334 | 1.28 | 0.747 | 0.590 | 0.946 | 25.3 | 5.4 | 41.0 |
| | D1 28+d | 363423 | 116 | 0.32 | 0.450 | 0.334 | 0.605 | 55.0 | 39.5 | 66.6 |
| | D2 0-13d | 100529 | 10 | 0.1 | 0.167 | 0.090 | 0.309 | 83.3 | 69.1 | 91.0 |
| | D2 14-83d | 91272 | 23 | 0.25 | 0.387 | 0.249 | 0.602 | 61.3 | 39.8 | 75.1 |
| | D2 84+d | 1936 | 2 | 1.03 | 4.500 | 1.019 | 19.885 | -350.0 | -1888.5 | -1.9 |

**Table S3**

| Vaccination Status | Nucleocapsid RNA target | | | | ORF1ab RNA target | | | |
|---|---|---|---|---|---|---|---|---|
| | n | mean Ct value | standard deviation | p value (two-tailed t test) | n | mean Ct value | standard deviation | p value (two-tailed t test) |
| Unvaccinated | 595 | 25.92 | 7.32 | - | 761 | 25.20 | 6.83 | - |
| Dose 1 28+d | 165 | 29.44 | 9.80 | <0.0001 | 143 | 26.65 | 8.76 | 0.0271 |
| Dose 2 14-83d | 58 | 30.86 | 7.07 | <0.0001 | 55 | 28.54 | 6.95 | 0.0005 |
| Dose 2 84+d | 310 | 25.82 | 7.51 | 0.8395 | 306 | 23.83 | 7.07 | 0.0033 |
| Dose 3 | 56 | 26.75 | 8.01 | 0.4209 | 53 | 24.14 | 7.22 | 0.2752 |
| Unvaccinated pre-Delta | 516 | 26.28 | 7.37 | - | 676 | 25.52 | 6.79 | - |
| Unvaccinated Delta | 79 | 23.58 | 6.59 | 0.0021 | 85 | 22.70 | 6.68 | 0.0003 |

**Table S4**

| **Residents** | | | | | | | | | | |
|---|---|---|---|---|---|---|---|---|---|---|
| **Prior SARS-CoV2 Status** | **Vaccination Status** | Person-days | Hospitalisation Events | Hospitalisation rate per 1000 person-days | aHR | L 95% CI | U 95% CI | VE | L 95% CI | U 95% CI |
| Unexposed | Unvaccinated | 544118 | 225 | 0.414 | 1 (ref) | | | | | |
| | AZ D1 0-27d | 151077 | 47 | 0.311 | 0.432 | 0.264 | 0.706 | 56.8 | 29.4 | 73.6 |
| | AZ D1 28+d | 280201 | 30 | 0.107 | 0.551 | 0.329 | 0.921 | 44.9 | 7.9 | 67.1 |
| | AZ D2 0-13d | 69998 | 4 | 0.057 | 0.615 | 0.141 | 2.694 | 38.5 | -169.4 | 85.9 |
| | AZ D2 14-83d | 366426 | 3 | 0.008 | 0.173 | 0.056 | 0.536 | 82.7 | 46.4 | 94.4 |
| | AZ D2 84+d | 583626 | 45 | 0.077 | 0.513 | 0.300 | 0.875 | 48.7 | 12.5 | 70.0 |
| | AZ D3 | 144716 | 5 | 0.035 | 0.067 | 0.026 | 0.172 | 93.3 | 82.8 | 97.4 |
| | mRNA D1 0-27d | 67230 | 20 | 0.297 | 0.376 | 0.189 | 0.749 | 62.4 | 25.1 | 81.1 |
| | mRNA D1 28+d | 121698 | 22 | 0.181 | 0.592 | 0.303 | 1.160 | 40.8 | -16.0 | 69.7 |
| | mRNA D2 0-13d | 30666 | 0 | 0.000 | 0.000 | . | . | 100.0 | | |
| | mRNA D2 14-83d | 161276 | 1 | 0.006 | 0.112 | 0.015 | 0.832 | 88.8 | 16.8 | 98.5 |
| | mRNA D2 84+d | 292101 | 14 | 0.048 | 0.349 | 0.184 | 0.664 | 65.1 | 33.6 | 81.6 |
| | mRNA D3 | 86703 | 7 | 0.081 | 0.164 | 0.073 | 0.366 | 83.6 | 63.4 | 92.7 |
| **Staff** | | | | | | | | | | |
| **Prior SARS-CoV2 Status** | **Vaccination Status** | Person-days | Hospitalisation Events | Hospitalisation rate per 1000 person-days | aHR | L 95% CI | U 95% CI | VE | L 95% CI | U 95% CI |
| Unexposed | Unvaccinated | 1046689 | 38 | 0.036 | 1 (ref) | | | | | |
| | AZ D1 0-27d | 146790 | 3 | 0.020 | 0.411 | 0.118 | 1.426 | 58.9 | -42.6 | 88.2 |
| | AZ D1 28+d | 296396 | 4 | 0.014 | 0.804 | 0.292 | 2.216 | 19.6 | -121.6 | 70.8 |
| | AZ D2 0-13d | 67057 | 0 | 0.000 | 0.000 | . | . | 100.0 | | |
| | AZ D2 14-83d | 347427 | 0 | 0.000 | 0.000 | . | . | 100.0 | | |
| | AZ D2 84+d | 518779 | 4 | 0.008 | 0.104 | 0.031 | 0.356 | 89.6 | 64.4 | 96.9 |

| | | | | | | | | | | |
|---|---|---|---|---|---|---|---|---|---|---|
| | AZ D3 | 81261 | 0 | 0.000 | 0.000 | . | . | 100.0 | | |
| | mRNA D1 0-27d | 169075 | 3 | 0.018 | 0.323 | 0.093 | 1.120 | 67.7 | -12.0 | 90.7 |
| | mRNA D1 28+d | 276699 | 0 | 0.000 | 0.000 | . | . | 100.0 | | |
| | mRNA D2 0-13d | 78805 | 0 | 0.000 | 0.000 | . | . | 100.0 | | |
| | mRNA D2 14-83d | 390613 | 0 | 0.000 | 0.000 | . | . | 100.0 | | |
| | mRNA D2 84+d | 633995 | 3 | 0.005 | 0.079 | 0.021 | 0.307 | 92.1 | 69.3 | 97.9 |
| | mRNA D3 | 155440 | 1 | 0.006 | 0.066 | 0.006 | 0.748 | 93.4 | 25.2 | 99.4 |

**Table S5**

| Residents | | | | | | | | | | |
|---|---|---|---|---|---|---|---|---|---|---|
| **Prior SARS-CoV2 Status** | **Vaccination Status** | Person-days | Deaths | Death rate per 1000 person-days | aHR | L 95% CI | U 95% CI | VE | L 95% CI | U 95% CI |
| Unexposed | Unvaccinated | 556900 | 293 | 0.53 | 1 (ref) | | | | | |
| | AZ D1 0-27d | 154656 | 58 | 0.38 | 0.202 | 0.127 | 0.321 | 79.8 | 67.9 | 87.3 |
| | AZ D1 28+d | 289546 | 36 | 0.12 | 0.473 | 0.268 | 0.836 | 52.7 | 16.4 | 73.2 |
| | AZ D2 0-13d | 72112 | 0 | 0.00 | 0.000 | . | . | 100.0 | | |
| | AZ D2 14-83d | 377974 | 2 | 0.01 | 0.083 | 0.020 | 0.349 | 91.7 | 65.1 | 98.0 |
| | AZ D2 84+d | 600790 | 33 | 0.05 | 0.389 | 0.205 | 0.738 | 61.1 | 26.2 | 79.5 |
| | AZ D3 | 148276 | 1 | 0.01 | 0.013 | 0.002 | 0.100 | 98.7 | 90.0 | 99.8 |
| | mRNA D1 0-27d | 67906 | 36 | 0.53 | 0.317 | 0.173 | 0.580 | 68.3 | 42.0 | 82.7 |
| | mRNA D1 28+d | 123441 | 31 | 0.25 | 0.414 | 0.230 | 0.747 | 58.6 | 25.3 | 77.0 |
| | mRNA D2 0-13d | 31095 | 2 | 0.06 | 0.423 | 0.096 | 1.875 | 57.7 | -87.5 | 90.4 |
| | mRNA D2 14-83d | 163794 | 0 | 0.00 | 0.000 | . | . | 100.0 | | |
| | mRNA D2 84+d | 296163 | 13 | 0.04 | 0.339 | 0.156 | 0.740 | 66.1 | 26.0 | 84.4 |
| | mRNA D3 | 87954 | 2 | 0.02 | 0.047 | 0.011 | 0.206 | 95.3 | 79.4 | 98.9 |
